# Vitamin D, acute respiratory infections, and Covid-19: the curse of small-size randomised trials. A critical review with meta-analysis of randomised trials

**DOI:** 10.1101/2024.04.26.24306354

**Authors:** Philippe Autier, Giulia Doi, Patrick Mullie, Patrick Vankrunkelsven, Oriana D’Ecclesiis, Sara Gandini

## Abstract

**Background:** Randomised trials conducted before 2021 indicated that vitamin D supplementation (VDS) was able to prevent severe COVID-19 and acute respiratory infections (ARI). However, these health benefits were not confirmed by larger randomised trials published after 2021.

**Objective:** To examine the characteristics of randomised trials on VDS to COVID-19 patients and admission to intensive care unit (ICU), and on VDS for the prevention of ARI.

**Method:** A systematic search retrieved randomised trials on VDS to COVID-19 patients and admission to ICU. Data on VDS and ARI were extracted from the meta-analysis of Jolliffe et al., 2021. The associations between VDS vs no VDS, and admission to ICU were evaluated using random effect models. Meta-analyses were done for all trials and by groups trial size. Publication bias was assessed using the LFK index (no bias if index between -1 and +1) and the Trim and Fill method.

**Results:** Nine trials on VDS for preventing admission to ICU were identified. The summary odds ratio (SOR) was 0.61 (95%CI: 0.39-0.95) for all trials, 0.34 (0.13-0.93) for trials including 50 to <106 patients and 0.88 (0.62-1.24) for trials including 106 to 548 patients (effect modification: p=0.04). The LFK index was -3.79, and after Trim and Fill, the SOR was 0.80 (0.40-1.61). The SOR for the 37 trials on VDS for ARI prevention was 0.92 (0.86-0.99) for all trials, 0.69 (0.57-0.83) for trials including 25 to <248 patients and 0.98 (0.94-1.03) for trials including 248 to 16,000 patients (effect modification p=0.0001). The LFK index was -3.11, and after Trim and Fill, the SOR was 0.96 (0.88-1.05).

**Conclusion:** Strong publication bias affected randomised trials on VDS for the prevention of severe COVID-19 and of ARI. Systematic reviews should beware of small-size randomised trials that generally exaggerate health benefits.

## Introduction

When the COVID-19 pandemic struck the World in 2019-2021, many thought that vitamin D supplementation (VDS) was an option for reducing the risk of COVID-19 and progression to severe disease [1–3]. The VDS option was nurtured by a wealth of data.

First, laboratory studies have documented the involvement of physiologically active forms of vitamin D in immunological mechanisms dealing with infectious agents [4, 5] [6, 7].

Second, the serum concentration of 25-hydroxyvitamin D (hereafter s-25OHD) is usually considered reflecting individual vitamin D status. Numerous observational studies done before and after the pandemic onset found first that patients with low s-25OHD were at higher risk of severe acute respiratory infections (ARI) and severe COVID-19 [8–10], and second that COVID-19 patients taking VDS had less severe disease course [11].

Third, a meta-analysis of aggregated data from 37 trials conducted between 2006 and 2019 found that VDS was associated with a 8% (95% confidence interval [CI]: 1% to 14%) reduction in the risk of ARI [12].

Fourth, randomised trials published in 2020 and 2021 obtained results compatible with a favourable effect of VDS on COVID-19 outcomes [13].

However, the promises of the VDS option were tempered by the publication in 2022 of large size randomised trials that found no effect of VDS on the incidence and severity of COVID-19 [14–16]. In addition, six Mendelian randomisation studies examined the risk of incidence and severity of COVID-19 in patients whose s-25OHD is lower than average because of their genetic background. Patients with naturally low s-25OHD had no greater risk of COVID-19 incidence or severity than patients with higher s-25OHD [17–22].

The disillusionment caused by Mendelian and large-size randomised trials called for a review of the thread of data which resulted in considering VDS as an option for the treatment of COVID-19. This article is a systematic review with meta-analysis of randomised trials on VDS for the prevention of severe COVID-19 in patients infected with the SARS-CoV-2 virus. This article also revisited the meta-analysis of randomised trials of VDS done before 2020 for the prevention of ARI [12].

## Methods

This review was not registered. A scoping review of the literature showed that the most robust outcome indicating severe COVID-19 disease was the admission to intensive care unit (ICU). Other possible outcomes were less robust (e.g., admission to hospital), or possibly influenced by subjectivity (e.g., self-reported dyspnoea, resolution of symptoms, duration of hospitalisation).

We performed a systematic search restricted to PubMed of randomised trials published since January 2000 on VDS and outcomes of COVID-19 patients aged 18 years or more. The search was done in two successive steps (S1 Text). We also searched references cited in review articles.

Studies were selected if (i) they reported on randomised trials that compared rates of admission to ICU or COVID-19 patients receiving VDS to patients not receiving VDS, (ii) VDS was the unique intervention, (iii) COVID-19 patients enrolled in trials were not taking VDS before trial enrolment, (iv) comparison groups included COVID-19 patients who did not receive any type of VDS during trial courses, and (v) published in English language. All forms of vitamin D were considered (ergocalciferol (vitamin D2), cholecalciferol (vitamin D3), calcifediol (25-hydroxycholecalciferol), calcitriol (1α,25-dihydroxycholecalciferol)).

Key study characteristics and data of selected studies were extracted by PA and PM, and independently verified by GD and OD. Extracted information were first author, publication year, patient numbers by randomization group, intervention, comparison groups (i.e., whether the VDS intervention was tested against a placebo, or no placebo), and numbers of patients with outcome by randomization group. Key data on patient numbers and outcomes (i.e., odds ratios [OR] and 95% CI) included in the meta-analysis of Jolliffe et al., 2021 [12] were extracted by PA and PM.

For trials in COVID-19 patients, risks of admission to ICU were computed as odds ratios with 95% CI following a per-protocol approach. Hence, statistical analyses were done using data from patients who took the VDS or the placebo (when applicable) according to trial protocols.

Standard errors (SE) of odds ratios (OR) were derived from the equation SQRT(1/a+1/b+1/c+1/d), where a and b are the numbers of patients admitted to ICU in the VDS and in the comparison group, respectively, and c and d are the numbers of patients not admitted to ICU in the VDS and in the comparison group, respectively.

For the meta-analysis on VDS of COVID-19 patients and admission in ICU, we used random-effect models to calculate summary odds ratios and 95% CI of admission to ICU of COVID-19 patients.

Random-effect models necessitate the estimation of the between-study variance τ^2^. Various methods exist for computing an estimator of the between-study variance τ^2^ and for estimating its confidence interval. Mathematical simulations have shown that the widely used Der Simonian-Laird estimator is known to be negatively biased in scenarios with small studies and in scenarios with a rare binary outcome [23].

Because most randomised trials found in the systematic search were of small and medium size, i.e., from about 40 to 400 patients enrolled, we opted to use the REML

(Restricted Maximum Likelihood) estimator, and the REML estimator combined with the HKSJ (Hartung-Knapp-Sidik-Jonkman) method for estimating the 95% CI [24, 25].

For the meta-analysis on VDS for ARI prevention [12], we computed pooled estimate applying the Der Simonian-Laird method in order to reproduce the original analysis.

Then for both meta-analyses, statistical heterogeneity was evaluated through the I^2^ index: a value <50% was considered indicating an absence of statistically significant between-study heterogeneity. To investigate the between-study heterogeneity and the stability of the pooled estimate, we carried out a sensitivity analysis by excluding from the meta-analysis one of the trials at a time.

We evaluated the presence of possible publication bias, using Egger’s regression test, and the Makaskill test. We also used the method proposed by Furuya-Kanamori et al. [26], which displays in a Doi plot the weight of each trial (i.e., the sample size of each trial relative to the size of all trials combined, expressed as a Z-score) against the natural logarithm of odds ratios reported by trials. The LFK index derived from the Doi plot provides a quantitative estimate of the asymmetry of odds ratios included in the meta-analysis. A LFK index of zero indicates an absence of asymmetry. Asymmetry, and thus the possibility of publication bias, is suggested by a LFK index less than -1 (studies not in favour of VDS less likely to be published) or greater than +1 (studies in favour of VDS less likely to be published).

We performed an exploratory analysis of publication bias using the Trim and Fill method [27], which provides summary odds ratios adjusted for publication bias.

We studied the effect of the sample size through a subgroup analysis, by dividing the trials based on their sample sizes (lower or greater than the median), and including in our statistical models the trial size as a possible effect modifier.

For trials on VDS in COVID-19 patients and admission to ICU, we assessed seven sources of bias following Cochrane’s rules. We attributed to each source a value +1 if the risk of bias was low (L), 0 if it was undetermined (U) and -1 if it was high (H). Then we created a summary score 1 which was the summation of values attributed to the seven sources. A summary score 2 was the summation of sources with low risk of bias (i.e., sources with +1 value).

To evaluate the influence of the risk of bias on results, we fitted meta-regression models to verify whether the quality scores were effect modifiers.

All reported p-values were two sided and p<0.05 was considered statistically significant. Meta-analyses were carried out by using the R-Studio software (R version 4.1.1).

## Results

### VDS to COVID-19 patients and admission to ICU

The literature search is depicted in Fig 1, S1 Table. Three articles [28–30] were found during hand search of review articles. After full article reading, nine articles [16, 28, 31–37] were selected for review and 14 articles were excluded. Reasons for exclusion were: high vs. low dose VDS [38–40], no specific numbers of patients who were admitted in ICU [14, 15, 30, 41–45], failure of randomization [29], and not a randomised trial [46, 47].

**Fig 1.**
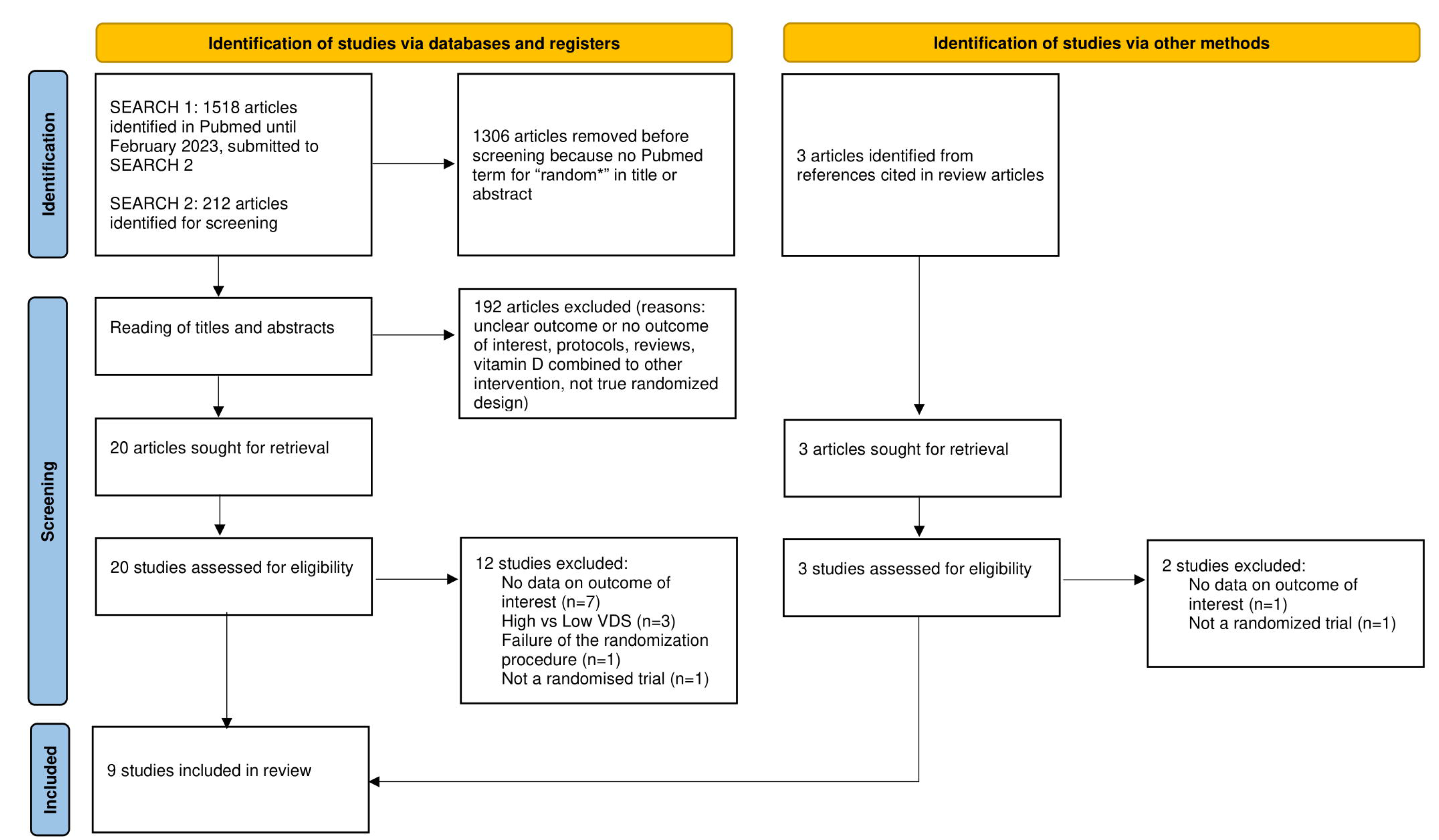
PRISMA 2020 literature search flow diagram (See S1 Text for terms used in Searches 1 and 2)

Characteristics of the nine selected randomised trials are summarized in Table 1. The vitamin D compound were cholecalciferol (5 trials), calcifediol (2 trials), or calcitriol (1 trial). VDS dosages and regimens were variable with daily intake in six trials and bolus administration in three trials. Four trials were placebo controlled and five trials were open-label. The numbers of COVID-19 patients enrolled in trials ranged from 50 to 548. The odds ratios for admission in ICU ranged from 0.02 to 1.06, with wide 95% CIs. The risk of bias analysis is detailed in S2 Table. Five trials had a low risk of bias, and four had a high risk of bias.

**Table 1.**
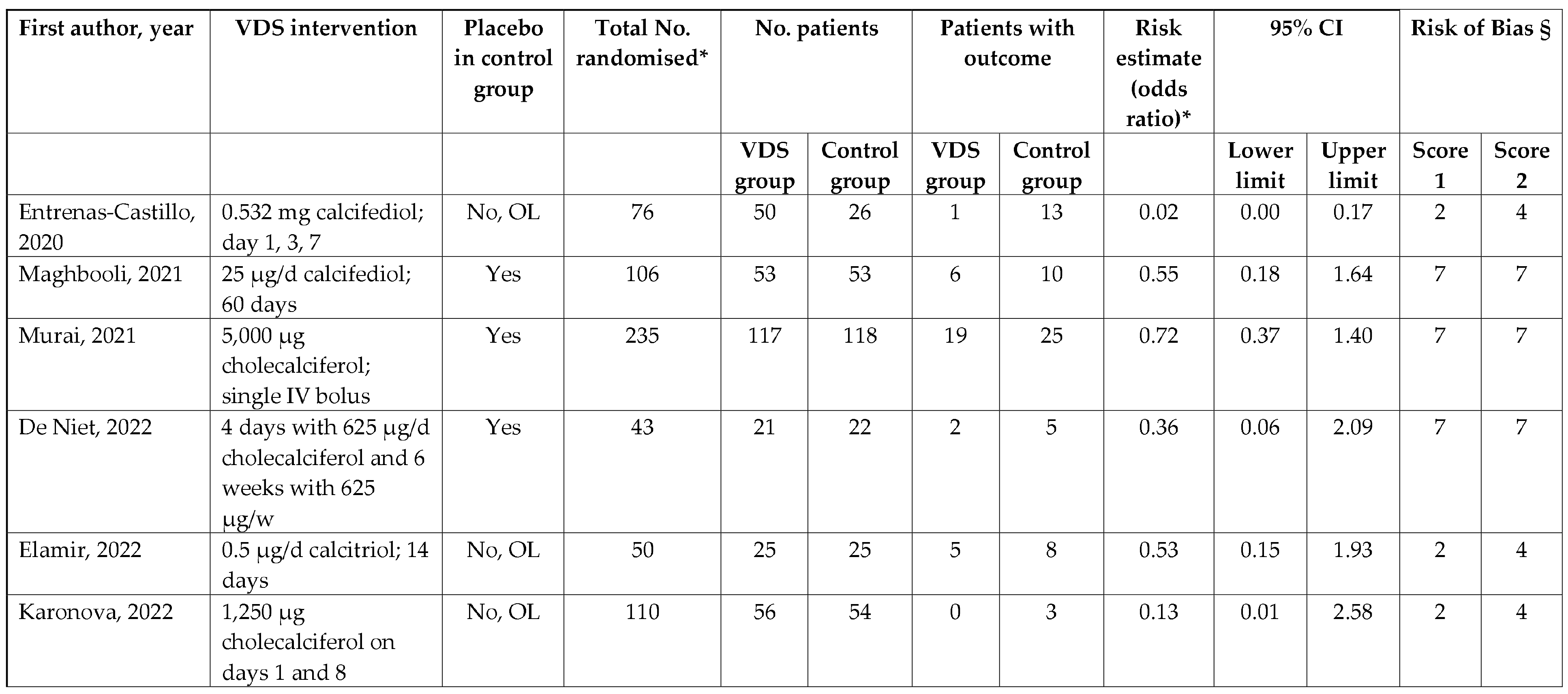
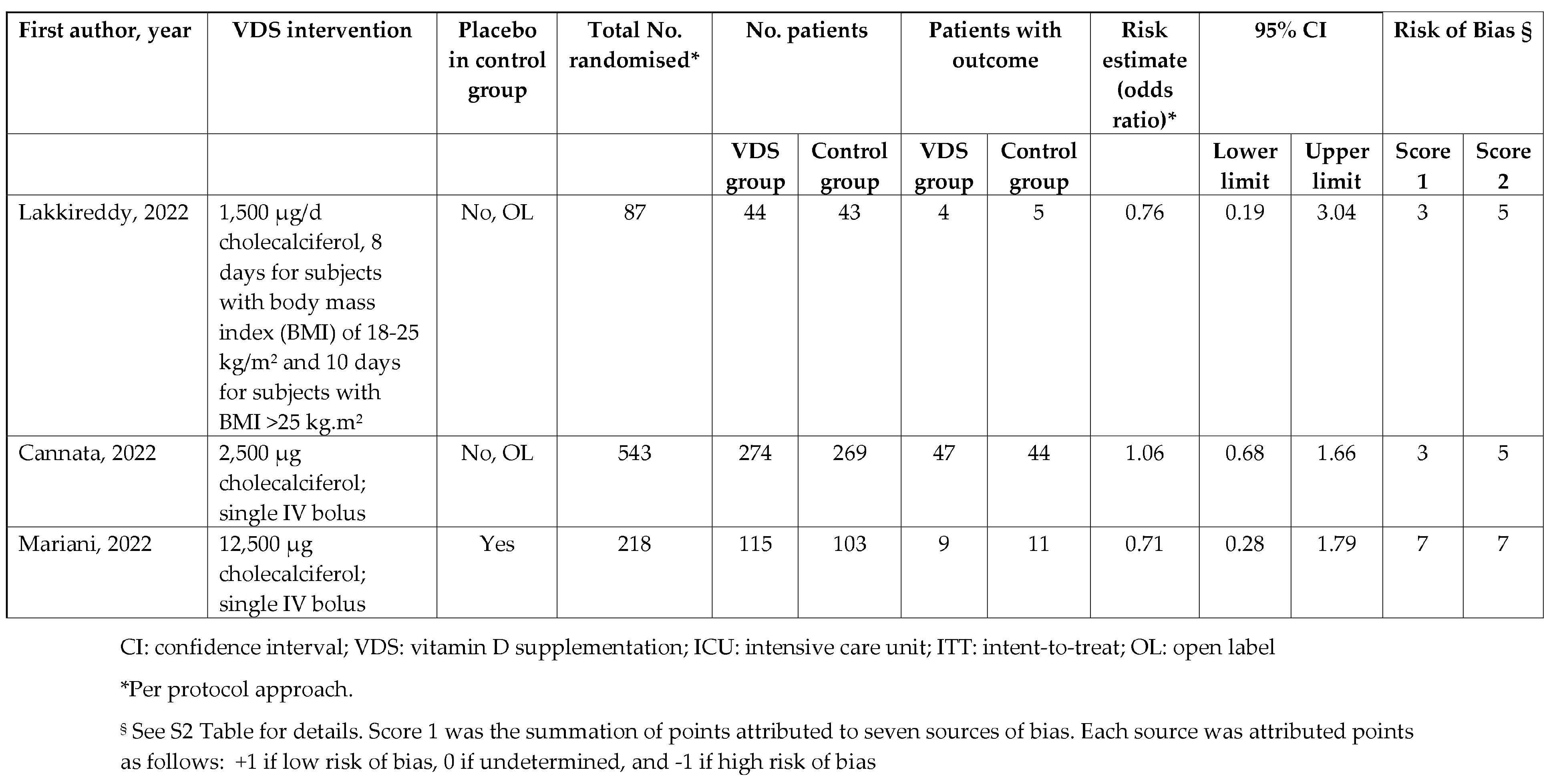
Randomised trials on vitamin D supplementation of COVID-19 patients and admission in ICU.

| First author, year | VDS intervention | Placebo in control group | Total No. randomised* | No. patients |  | Patients with outcome |  | Risk estimate (odds ratio)* | 95% CI |  | Risk of Bias § |  |
| --- | --- | --- | --- | --- | --- | --- | --- | --- | --- | --- | --- | --- |
|  |  |  |  | VDS group | Control group | VDS group | Control group |  | Lower limit | Upper limit | Score 1 | Score 2 |
| Entrenas-Castillo, 2020 | 0.532 mg calcifediol; day 1, 3, 7 | No, OL | 76 | 50 | 26 | 1 | 13 | 0.02 | 0.00 | 0.17 | 2 | 4 |
| Maghbooli, 2021 | 25 µg/d calcifediol; 60 days | Yes | 106 | 53 | 53 | 6 | 10 | 0.55 | 0.18 | 1.64 | 7 | 7 |
| Murai, 2021 | 5,000 µg cholecalciferol; single IV bolus | Yes | 235 | 117 | 118 | 19 | 25 | 0.72 | 0.37 | 1.40 | 7 | 7 |
| De Niet, 2022 | 4 days with 625 µg/d cholecalciferol and 6 weeks with 625 µg/w | Yes | 43 | 21 | 22 | 2 | 5 | 0.36 | 0.06 | 2.09 | 7 | 7 |
| Elamir, 2022 | 0.5 µg/d calcitriol; 14 days | No, OL | 50 | 25 | 25 | 5 | 8 | 0.53 | 0.15 | 1.93 | 2 | 4 |
| Karonova, 2022 | 1,250 µg cholecalciferol on days 1 and 8 | No, OL | 110 | 56 | 54 | 0 | 3 | 0.13 | 0.01 | 2.58 | 2 | 4 |
|  |  |  |  | VDS group | Control group | VDS group | Control group |  | Lower limit | Upper limit | Score 1 | Score 2 |
| Lakkireddy, 2022 | 1,500 µg/d cholecalciferol, 8 days for subjects with body mass index (BMI) of 18-25 kg/m<sup>2</sup> and 10 days for subjects with BMI >25 kg.m<sup>2</sup> | No, OL | 87 | 44 | 43 | 4 | 5 | 0.76 | 0.19 | 3.04 | 3 | 5 |
| Cannata, 2022 | 2,500 µg cholecalciferol; single IV bolus | No, OL | 543 | 274 | 269 | 47 | 44 | 1.06 | 0.68 | 1.66 | 3 | 5 |
| Mariani, 2022 | 12,500 µg cholecalciferol; single IV bolus | Yes | 218 | 115 | 103 | 9 | 11 | 0.71 | 0.28 | 1.79 | 7 | 7 |

CI: confidence interval; VDS: vitamin D supplementation; ICU: intensive care unit; ITT: intent-to-treat; OL: open label
\*Per protocol approach.
§ See S2 Table for details. Score 1 was the summation of points attributed to seven sources of bias. Each source was attributed points as follows: +1 if low risk of bias, 0 if undetermined, and -1 if high risk of bias

The meta-analysis of the nine trials based on the RMLE estimator resulted in a summary odds ratio of 0.61 (95% CI: 0.39-0.95) (Fig 2, Table 2), with low heterogeneity (I^2^=35%), suggesting that overall COVID-19 patients taking VDS would have a substantially decreased risk of admission to ICU.

**Fig 2.**
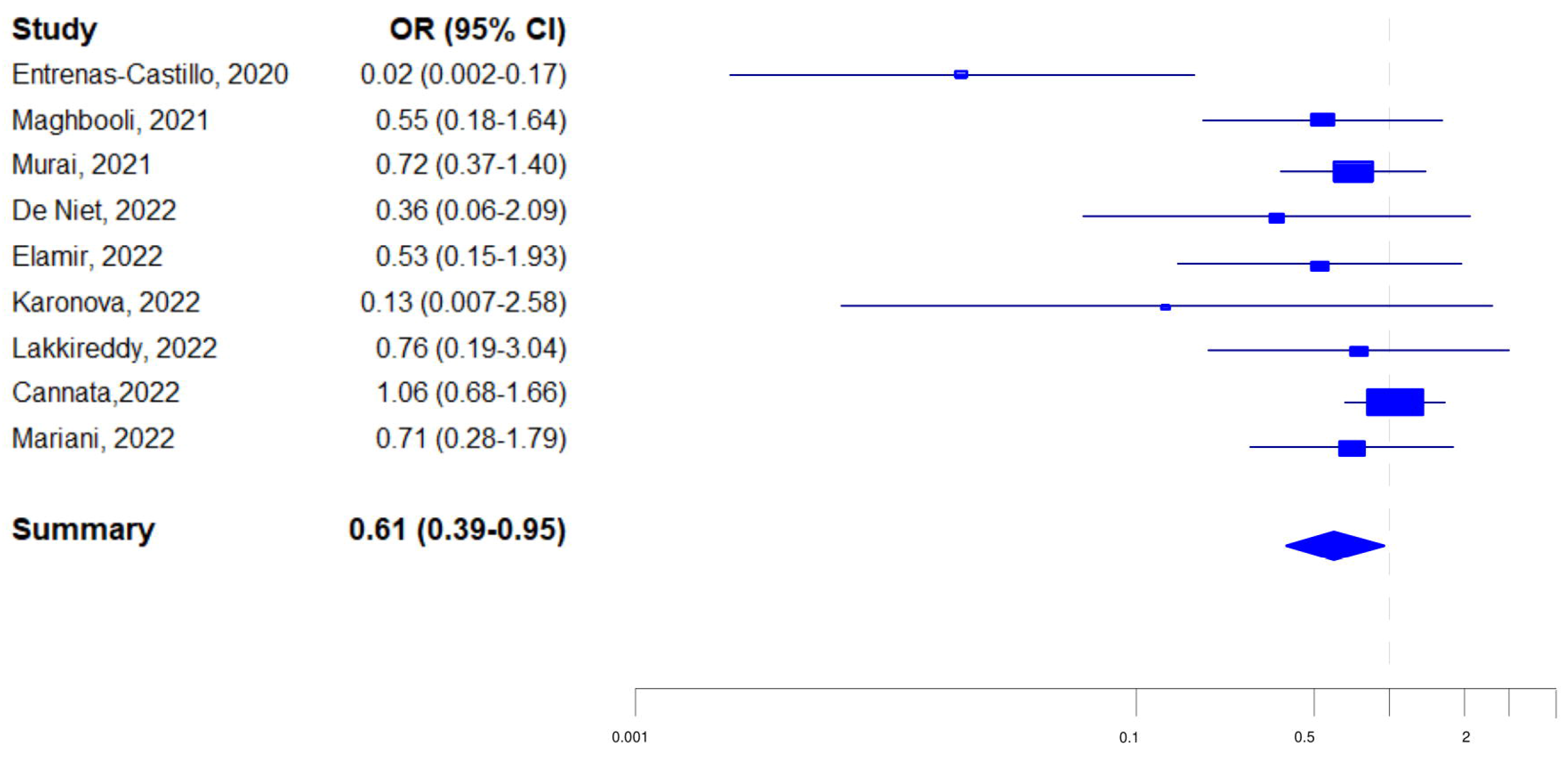
Forest plot of randomised trials on VDS of COVID-19 patients and admission to ICU.

**Table 2.** Meta-analysis of randomised trials on VDS of COVID-19 patients and admission in ICU.

|  |  |  |  |
| --- | --- | --- | --- |
| **Restricted maximum likelihood estimator** |  |  |  |
| 9 trials |  |  |  |
| Summary risk estimate | SOR (95% CI) | 0.61 | (0.39-0.95)* |
| Heterogeneity | I<sup>2</sup> | 35% |  |
| Publication bias | Egger test | p=0.001 |  |
|  | Modified Macaskill test | p=0.03 |  |
| Summary risk estimate after Trim&Fill | OR (95% CI) | 0.80 | (0.40-1.61) |
| 8 trials without Entrenas-Castillo et al, 2020 |  |  |  |
| Summary risk estimate | SOR (95% CI) | 0.78 | (0.57-1.08) |
| Heterogeneity | I<sup>2</sup> | 4% |  |
| 5 trials which included less than 106 patients |  |  |  |
| Summary risk estimate | SOR (95% CI) | 0.34 | (0.13-0.93) |
| 4 trials which included 106 patients or more |  |  |  |
| Summary risk estimate | SOR (95% CI) | 0.88 | (0.62-1.24) |
| Interaction between randomisation group and trial size § |  |  | p=0.04 |
| **Restricted Maximum Likelihood Estimator plus Hartung-Knapp-Sidik-Jonkman for 95% CI** |  |  |  |
| 9 trials |  |  |  |
| Summary risk estimate | SOR (95% CI) | 0.61 | (0.32-1.16) |
| Heterogeneity | I<sup>2</sup> | 35% |  |
| Publication bias | Egger test | p=0.0086 |  |
|  | Modified Macaskill test | p=0.1195 |  |
| Summary risk estimate after Trim&Fill | OR (95% CI) | 0.80 | (0.33-1.97) |
| 8 trials without Entrenas-Castillo et al, 2020 |  |  |  |
| Summary risk estimate | SOR (95% CI) | 0.78 | (0.58-1.06) |
| Heterogeneity | I<sup>2</sup> | 3.7% |  |
| 5 trials which included less than 106 patients |  |  |  |
| Summary risk estimate | SOR (95% CI) | 0.34 | (0.07-1.72) |
| 4 trials which included 106 patients or more |  |  |  |
| Summary risk estimate | SOR (95% CI) | 0.88 | (0.51-1.50) |
| Interaction between randomisation group and trial size § |  |  | p=0.16 |

ICU: intensive care unit; OR: odds ratio; SOR: summary odds ratio; VDS: vitamin D supplementation
\* 0.05>p>0.001
§ Interaction term combining randomisation group (VDS vs comparison) to trial size (<106 vs ≥106 patients)

The Egger’ and the modified Macaskill tests indicated significant publication bias (Table 2). The display of odds ratios from the nine trials in a Doi plot (Fig 3a) showed a distribution markedly stretched to the left, witnessing a negative publication bias, i.e., trials not favouring a reduction of admissions to ICU associated with VDS were less likely to be published than trials favouring a reduction of admissions to ICU associated with VDS. Of note, the smaller the trial size (i.e., the higher the Z-score), the greater the risk reductions, i.e., ln(OR) moving towards negative values far from zero.

**Fig 3.**
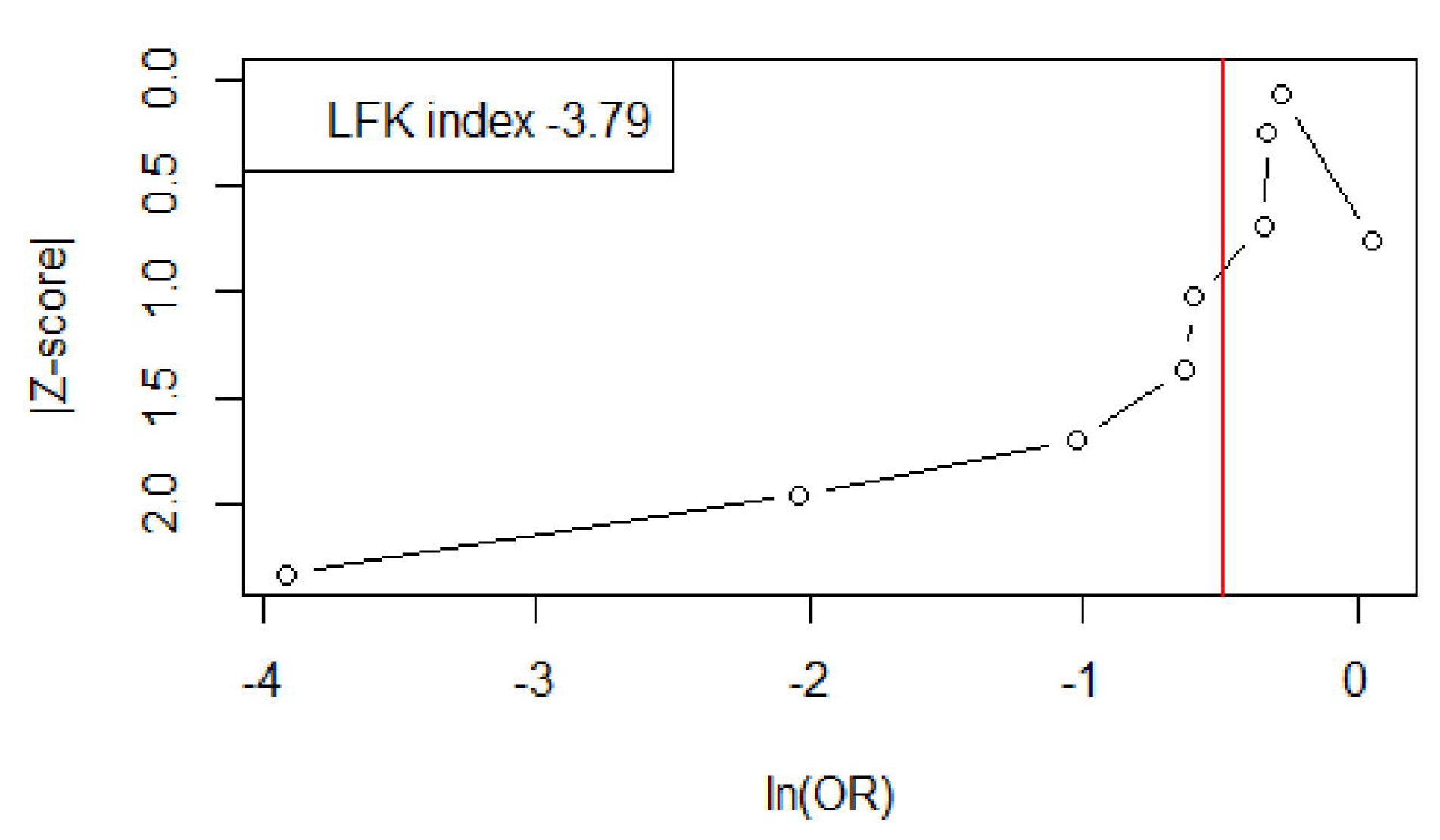
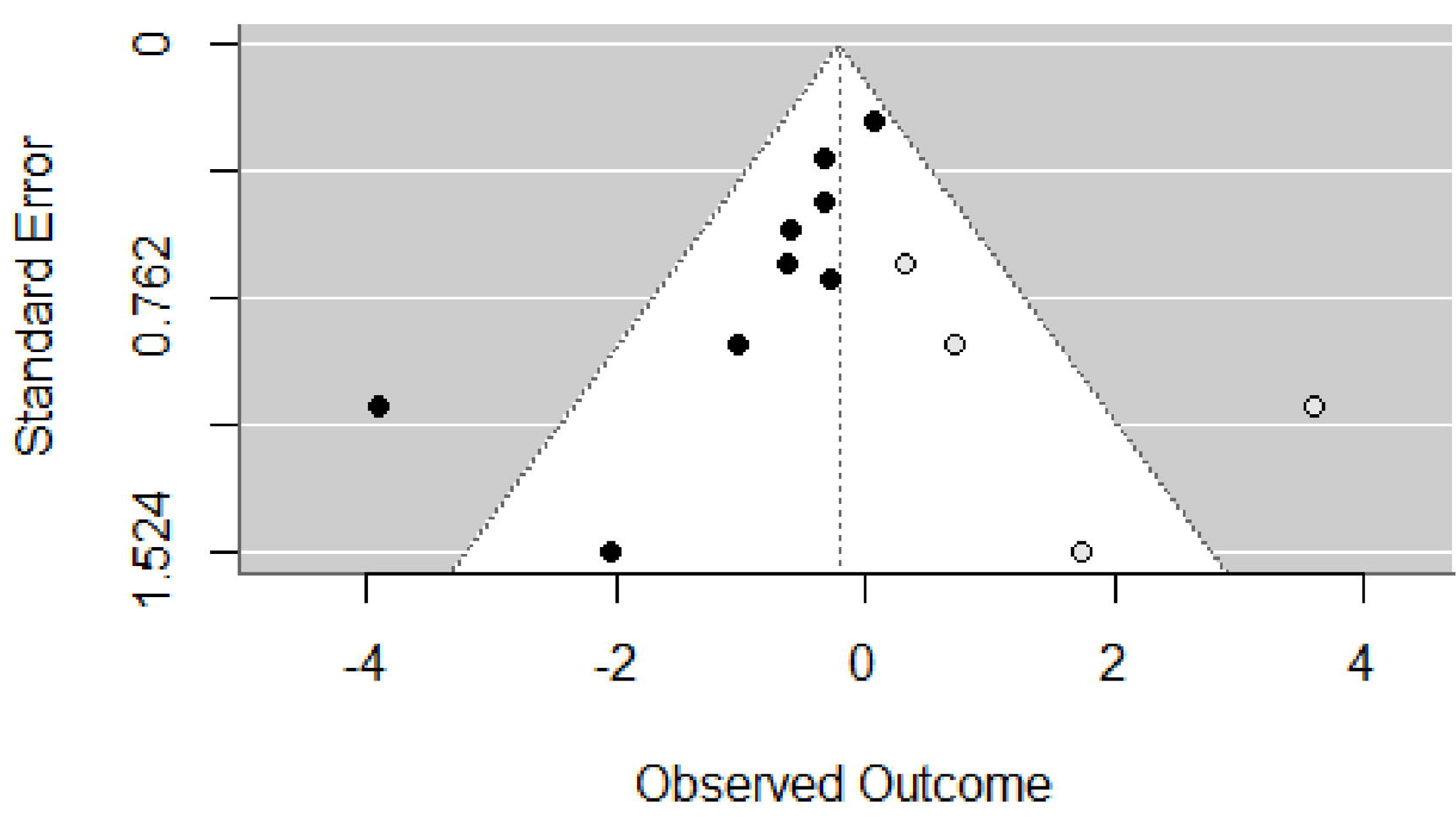
Doi plot (Fig 3a) and funnel plot after Trim and Fill correction (Fig 3b) for randomised trials on VDS and admission to ICU

The LFK index of -3.79, well below -1.0, quantifies the strong imbalance by which trials in favour of VDS had a much greater probability to be published than trials in favour of VDS.

The Trim and Fill method resulted in the addition of four open circles on the funnel plot (Fig 3b). These open circles are imputations of results of four hypothetical smaller-size trials not in favour of VDS that should have been published for completely excluding the possibility of publication bias. After application of the Trim and Fill method, the summary odds ratio increased to 0.80 (95% CI: 0.40-1.61), which was no longer significant.

Exclusion of one trial at a time from the meta-analysis showed that exclusion of Entrenas-Castillo et al, 2020 [37] of the meta-analysis ended up in a non-significant summary odds ratio of 0.78, with a drastic drop in heterogeneity between results of the eight other trials (I^2^=4%).

After stratification of trials according to numbers of patients enrolled (Table 2), the summary odds ratios were 0.34 (95% CI: 0.13-0.93) for the five trials including 50 to 105 COVID-19 patients, and 0.88 (95% CI: 0.62-1.24) for the four trials including 106 to 548 patients. The trial size had a significant modification effect on summary odds ratios (p-value = 0.04).

A meta-regression model showed that the two risk of bias scores (Table 1) did not significantly modify meta-analysis results: p-values were 0.46 for score 1 and 0.34 for score 2.

Use of the RMLE estimator with Hartung-Knapp-Sidik-Jonkman (HKSJ) method for the 95% CI resulted in a summary odds ratio of 0.61 (95% CI: 0.32-1.16). If the summary odds ratio remained the same, the 95% CI was wider, from 0.56 (=0.95-0.39) when the RMLE estimator only was used to 0.72 (=1.16-9.32) when the RMLE estimator and the HKSJ methods were used. Because of the wider 95% CI, the risk of admission to ICU was no longer significant. All other meta-analysis results remained equivalent or close

to results when the RMLE estimator only was used, but owing to the larger confidence interval, the effect modification with trial size was no longer significant.

### VDS for ARI prevention

The 37 trials on VDS for the prevention of ARI included in the publication of Jolliffe et al, 2021 [12] enrolled 25 to 16,000 subjects (median of 247). The meta-analysis of the 37 trials resulted in a summary odds ratio of 0.92 (95% CI: 0.86-0.99), suggesting a beneficial effect of VDS on the risk of ARI. But the Egger and the modified Macaskill tests provided evidence for significant publication bias (Table 3).

**Table 3.** Meta-analysis of randomised trials on VDS of the prevention of ARI and admission in ICU.

|  |  |  |  |
| --- | --- | --- | --- |
| **Restricted Maximum Likelihood Estimator** |  |  |  |
| 37 trials |  |  |  |
| SRE | OR (95% CI) | 0.92 | (0.86-0.99)* |
| Heterogeneity | I<sup>2</sup> | 36% |  |
| Publication bias | Egger test | p=0.007 |  |
|  | Modified Macaskill test | p=0.0004 |  |
| After Trim&Fill | OR (95% CI) | 0.96 | (0.88-1.05) |
| 19 trials which included less than 248 patients |  |  |  |
| SRE | OR (95% CI) | 0.69 | (0.57-0.83) |
| 18 trials which included 248 patients or more |  |  |  |
| SRE | OR (95% CI) | 0.98 | (0.94-1.03) |
| Interaction between randomisation group and trial size § |  | p=0.0001 |  |

ARI: acute respiratory infection; VDS: vitamin D supplementation; OR: odds ratio.
§ Interaction term combining randomisation group (VDS vs comparison) to trial size (<248 vs ≥248 patients)

The display of odds ratios from the 37 trials in a Doi plot (Fig 4a) showed a distribution markedly stretched to the left, witnessing a negative publication bias, i.e., trials not favouring a reduction of ARI associated with VDS were less likely to be published than trials favouring a reduction of ARI associated with VDS. The smaller the trial size (i.e., the higher the Z-Score), the greater the reduction of the risk of ARI, i.e., ln(OR) moving towards negative values far from zero.

**Fig 4.**
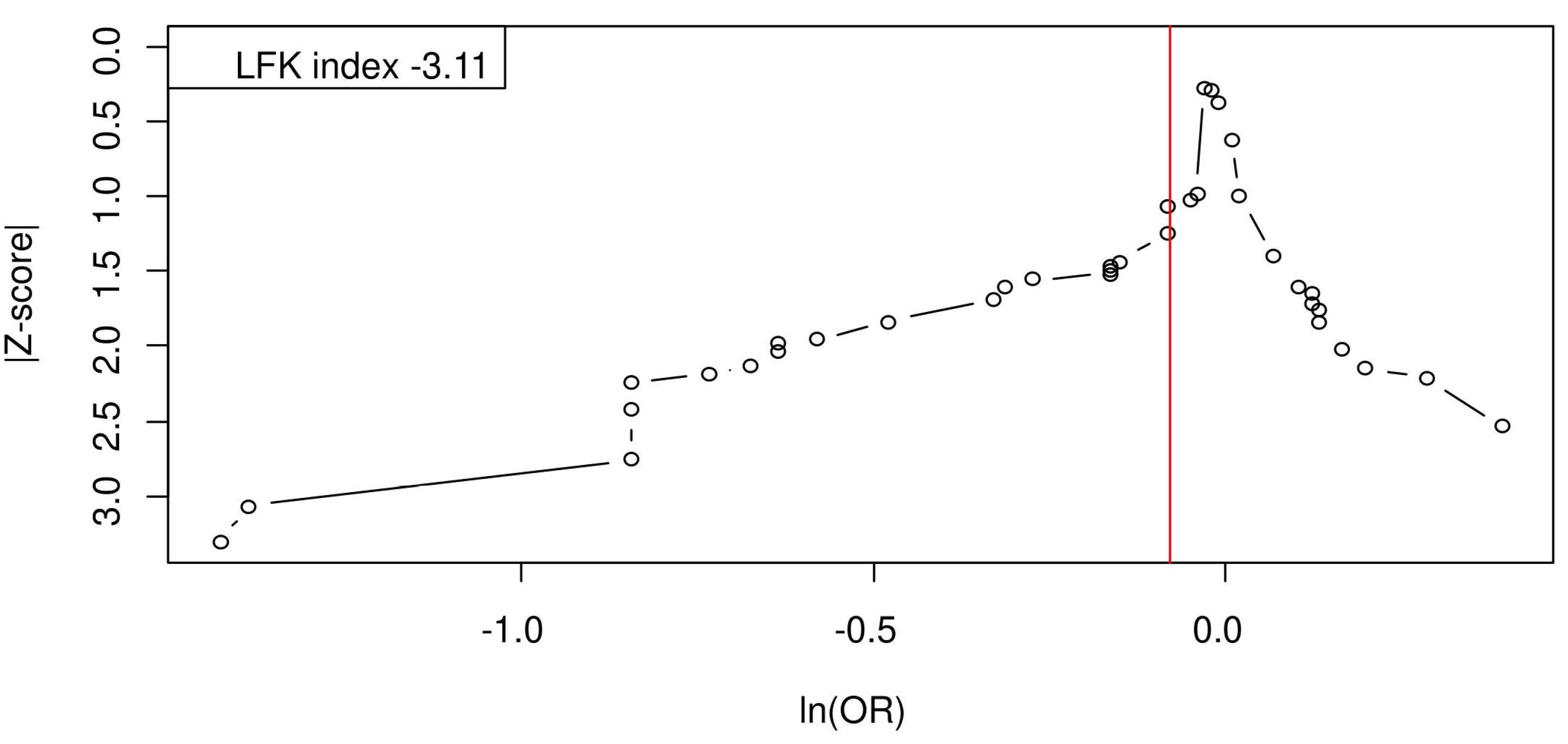
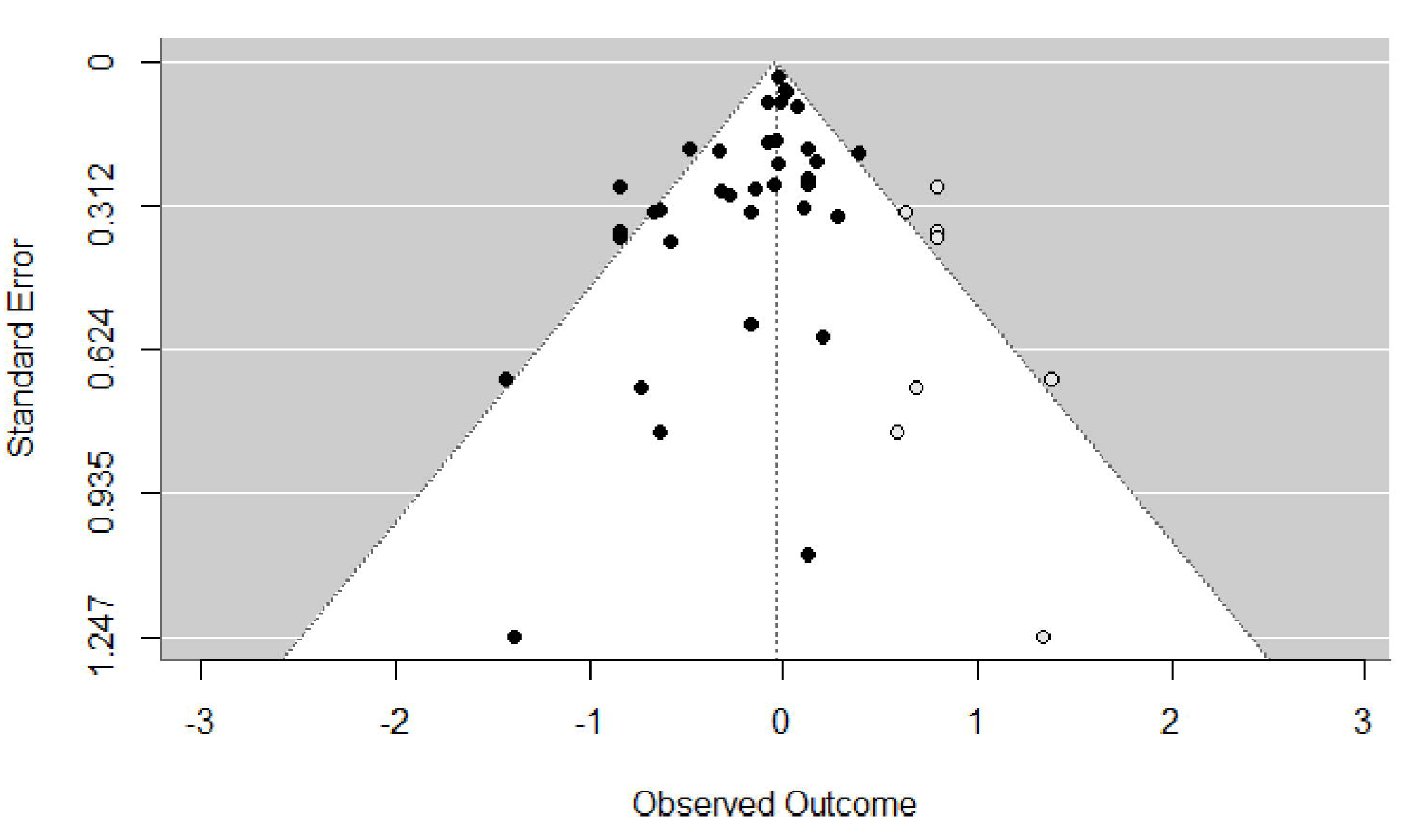
Doi plot (Fig 4a) and funnel plot after Trim and Fill correction (Fig 4b) for randomised trials on VDS and occurrence of ARI.

The LFK index of -3.11, well below -1.0, quantifies the strong imbalance by which trials in favour of VDS had a much greater probability to be published than trials not in favour of VDS.

The Trim and Fill method resulted in the addition of eight open-circle points on the funnel plot (Fig 4b). These open-circle points are imputed results of hypothetical smaller-size trials not in favour of VDS that should have been published for completely excluding the possibility of publication bias. After application of the Trim and Fill method, the summary odds ratio increased to 0.96 (95% CI: 0.88-1.95), which was no longer significant.

The meta-analysis of the 19 trials including 25 to 247 patients obtained a summary odds ratio of 0.69 (95% CI: 0.57-0.83) suggesting marked protective effect of VDS. Instead, the meta-analysis of the 18 trials including 250 to 16,000 patients or more resulted in a summary odds ratio of 0.98 (95% CI: 0.94-1.03) indicating no protective effect of VDS. The trial size had a significant modification effect on summary odds ratios (p-value = 0.0001).

## Discussion

Our review on VDS for COVID-19 patients and the risk of admission to ICU, and on VDS for the prevention of ARI shows that the majority of small-size randomised trials obtained results in sharp contrast with results of medium-to-large-size randomised trials. Results of medium-size randomised trials in the COVID-19 setting are in agreement with the six aforementioned Mendelian randomization studies that examined the risk of incidence and severity of COVID-19 according to s-25OHD. They

also agree with the negative results of large-size randomised trials conducted before 2020 on high-dose VDS for critically ill, vitamin D deficient patients admitted in intensive care unit [48, 49].

Hence publication bias was probably present, meaning that a number of small-size trials that obtained odds ratio not in favour of VDS were not published because either authors did not consider that their work was worth being submitted to medical journals, or because submissions were turned down by medical journals.

When the COVID-19 pandemic expanded, many new open-source resources were created for rapid dissemination of research findings, with the consequence of more lenience on the quality of research [50].

Jolliffe et al., 2021 [12] considered that virtually all 37 trials were at low risk of bias, which is surprising in view of the contrasting results between small-size and large-size trials. Although in the original publication a funnel plot highlighted a left-sided asymmetry with Egger’s test having a p-value of 0.007, the authors considered that these results “might reflect heterogeneity of effect across trials, or publication bias arising from omission of small trials showing non-protective effects of vitamin D supplementation from the meta-analysis”.

The interpretation of small-size trial effects is notoriously tricky. Overrepresentation of small-size trials with substantial effect on outcome can be due to publication bias and to

other reasons like poorer quality of small-size trials compared to large-size trials, or selective reporting of results [51–54].

Systematic reviews with meta-analyses of randomised trials are considered the optimal methodology for evaluating treatment efficacy. However, our study shows that, regardless of being the result of publication bias or poor-quality research, small-size randomised trials can lead to overestimate the efficacy of interventions [54, 55]. The undesirable effects of small-size trials have been reported for a variety of therapeutic areas [56], including critical care [57].

## Study limitations

Our study has several limitations. The study unfolded with the uncovering of the effects of trial sizes on the associations between VDS and acute respiratory infections or COVID-19 severity. In this regard, the study did not follow a protocol written before looking at the data. We reused data from Jolliffe et al, 2021 without a literature search for more recent publication on randomised trials testing VDS for acute respiratory infections. The literature search on VDS for COVID-19 was restricted to the Pubmed and reference lists of review articles published in 2022. However, we believe that more extensive literature searches were not likely to find other publications relevant to this work.

## Low vitamin D status and reverse causation

The negative results of medium-size trials on VDS are in striking contrast with the common observation that low s-25OHD is associated with more severe COVID-19 and death. This contrast underscores that low s-25OHD would have no detrimental role in COVID-19 but would rather be the consequence of two sets of factors (i.e., reverse causation). First, s-25OHD seems to behave like a negative surface reactant whose serum concentration fall during inflammatory states, just like albumin, transferrin and other compounds [41] [58] [59–61]. Second, patient’s characteristics and medical history known to precipitate severe COVID-19, like age, diabetes, obesity, frailty, cardiovascular conditions, are also known risk factors for low s-25OHD [62].

The associations between low s-25OHD and COVID-19 severity echoes the many observational studies showing that patients with low s-25OHD are at higher risk of acute and chronic disease, including cardiovascular diseases, diabetes, and fractures, and of premature death [62, 63]. However, randomised trials and their meta-analyses have failed to confirm that VDS could prevent or treat any of these diseases and prolong life-expectancy [64–67].

## Conclusions

Our review showed that the alleged protection of VDS against severe COVID-19 was the consequence of small-size randomised trials published before and in the early stages of the COVID-19 pandemic. If in 2022, it was recognised that VDS brought no benefit to COVID-19 patients [68, 69], mechanistic data, observational studies and small-size randomised trials done before have contributed to treating many COVID-19 patients with VDS. Even if VDS does not cause serious side effect, the same thread of laboratory and observational studies, and small size randomised trials could lead to the adoption of poorly effective but unsafe preventive or therapeutic interventions, until larger randomised trials would discourage their use.

## Supporting information

S1 Text. Keywords used for literature search in Pubmed and Prisma Flow Chart. S1 Table. PRISMA 2020 checklist

S2 Table. Risk of bias in randomised trials on VDS of COVID-19 patients and admission to ICU

## Data Availability

All relevant data are within the manuscript and its Supporting Information files. Articles from which data were extracted are publicly available and cited in references.

## References

1. Chakhtoura M, Napoli N, El Hajj Fuleihan G. Commentary: Myths and facts on vitamin D amidst the COVID-19 pandemic. Metabolism. 2020;109:154276. doi: 10.1016/j.metabol.2020.154276

2. Bergman P. The link between vitamin D and COVID-19: distinguishing facts from fiction. J Intern Med. 2021;289(1):131–3. doi: 10.1111/joim.13158

3. Martineau AR, Forouhi NG. Vitamin D for COVID-19: a case to answer? Lancet Diabetes Endocrinol. 2020;8(9):735–6. doi: 10.1016/s2213-8587(20)30268-0

4. Liu PT, Stenger S, Li H, Wenzel L, Tan BH, Krutzik SR, et al. Toll-like receptor triggering of a vitamin D-mediated human antimicrobial response. Science. 2006;311(5768):1770-3. doi: 10.1126/science.1123933

5. Sjoding MW, Luo K, Miller MA, Iwashyna TJ. When do confounding by indication and inadequate risk adjustment bias critical care studies? A simulation study. Crit Care. 2015;19(1):195. doi: 10.1186/s13054-015-0923-8

6. Lopez DV, Al-Jaberi FAH, Woetmann A, Ødum N, Bonefeld CM, Kongsbak-Wismann M, et al. Macrophages Control the Bioavailability of Vitamin D and Vitamin D-Regulated T Cell Responses. Front Immunol. 2021;12:722806. doi: 10.3389/fimmu.2021.722806

7. Chauss D, Freiwald T, McGregor R, Yan B, Wang L, Nova-Lamperti E, et al. Autocrine vitamin D signaling switches off pro-inflammatory programs of T(H)1 cells. Nat Immunol. 2022;23(1):62–74. doi: 10.1038/s41590-021-01080-3

8. Li-Ng M, Aloia JF, Pollack S, Cunha BA, Mikhail M, Yeh J, et al. A randomized controlled trial of vitamin D3 supplementation for the prevention of symptomatic upper respiratory tract infections. Epidemiol Infect. 2009;137(10):1396–404. doi: 10.1017/S0950268809002404

9. Pham H, Rahman A, Majidi A, Waterhouse M, Neale RE. Acute Respiratory Tract Infection and 25-Hydroxyvitamin D Concentration: A Systematic Review and Meta-Analysis. Int J Environ Res Public Health. 2019;16(17). doi: 10.3390/ijerph16173020

10. Dissanayake HA, de Silva NL, Sumanatilleke M, de Silva SDN, Gamage KKK, Dematapitiya C, et al. Prognostic and Therapeutic Role of Vitamin D in COVID-19: Systematic Review and Meta-analysis. J Clin Endocrinol Metab. 2022;107(5):1484–502. doi: 10.1210/clinem/dgab892

11. D’Ecclesiis O, Gavioli C, Martinoli C, Raimondi S, Chiocca S, Miccolo C, et al. Vitamin D and SARS-CoV2 infection, severity and mortality: A systematic review and meta-analysis. PLoS One. 2022;17(7):e0268396. doi: 10.1371/journal.pone.0268396

12. Jolliffe DA, Camargo CA, Jr., Sluyter JD, Aglipay M, Aloia JF, Ganmaa D, et al. Vitamin D supplementation to prevent acute respiratory infections: a systematic review and meta-analysis of aggregate data from randomised controlled trials. Lancet Diabetes Endocrinol. 2021;9(5):276–92. doi: 10.1016/S2213-8587(21)00051-6

13. Kummel LS, Krumbein H, Fragkou PC, Hunerbein BL, Reiter R, Papathanasiou KA, et al. Vitamin D supplementation for the treatment of COVID-19: A systematic review and meta-analysis of randomized controlled trials. Front Immunol. 2022;13:1023903. doi: 10.3389/fimmu.2022.1023903

14. Jolliffe DA, Holt H, Greenig M, Talaei M, Perdek N, Pfeffer P, et al. Effect of a test-and-treat approach to vitamin D supplementation on risk of all cause acute respiratory tract infection and covid-19: phase 3 randomised controlled trial (CORONAVIT). Bmj. 2022;378:e071230. doi: 10.1136/bmj-2022-071230

15. Brunvoll SH, Nygaard AB, Ellingjord-Dale M, Holland P, Istre MS, Kalleberg KT, et al. Prevention of covid-19 and other acute respiratory infections with cod liver oil supplementation, a low dose vitamin D supplement: quadruple blinded, randomised placebo controlled trial. Bmj. 2022;378:e071245. doi: 10.1136/bmj-2022-071245

16. Cannata-Andia JB, Diaz-Sottolano A, Fernandez P, Palomo-Antequera C, Herrero-Puente P, Mouzo R, et al. A single-oral bolus of 100,000 IU of cholecalciferol at hospital admission did not improve outcomes in the COVID-19 disease: the COVID-VIT-D-a randomised multicentre international clinical trial. BMC Med. 2022;20(1):83. doi: 10.1186/s12916-022-02290-8

17. Ma H, Zhou T, Heianza Y, Qi L. Habitual use of vitamin D supplements and risk of coronavirus disease 2019 (COVID-19) infection: a prospective study in UK Biobank. Am J Clin Nutr. 2021;113(5):1275–81. doi: 10.1093/ajcn/nqaa381

18. Butler-Laporte G, Nakanishi T, Mooser V, Morrison DR, Abdullah T, Adeleye O, et al. Vitamin D and COVID-19 susceptibility and severity in the COVID-19 Host Genetics Initiative: A Mendelian randomization study. PLoS Med. 2021;18(6):e1003605. doi: 10.1371/journal.pmed.1003605

19. Patchen BK, Clark AG, Gaddis N, Hancock DB, Cassano PA. Genetically predicted serum vitamin D and COVID-19: a Mendelian randomisation study. BMJ Nutr Prev Health. 2021;4(1):213–25. doi: 10.1136/bmjnph-2021-000255

20. Cui Z, Tian Y. Using genetic variants to evaluate the causal effect of serum vitamin D concentration on COVID-19 susceptibility, severity and hospitalization traits: a Mendelian randomization study. J Transl Med. 2021;19(1):300. doi: 10.1186/s12967-021-02973-5

21. Li X, van Geffen J, van Weele M, Zhang X, He Y, Meng X, et al. An observational and Mendelian randomisation study on vitamin D and COVID-19 risk in UK Biobank. Sci Rep. 2021;11(1):18262. doi: 10.1038/s41598-021-97679-5

22. Amin HA, Drenos F. No evidence that vitamin D is able to prevent or affect the severity of COVID-19 in individuals with European ancestry: a Mendelian randomisation study of open data. BMJ Nutr Prev Health. 2021;4(1):42–8. doi: 10.1136/bmjnph-2020-000151

23. Cornell JE, Mulrow CD, Localio R, Stack CB, Meibohm AR, Guallar E, et al. Random-effects meta-analysis of inconsistent effects: a time for change. Ann Intern Med. 2014;160(4):267–70. doi: 10.7326/M13-2886

24. Langan D, Higgins JPT, Jackson D, Bowden J, Veroniki AA, Kontopantelis E, et al. A comparison of heterogeneity variance estimators in simulated random-effects meta-analyses. Res Synth Methods. 2019;10(1):83–98. doi: 10.1002/jrsm.1316

25. Veroniki AA, Jackson D, Viechtbauer W, Bender R, Bowden J, Knapp G, et al. Methods to estimate the between-study variance and its uncertainty in meta-analysis. Res Synth Methods. 2016;7(1):55–79. doi: 10.1002/jrsm.1164

26. Furuya-Kanamori L, Barendregt JJ, Doi SAR. A new improved graphical and quantitative method for detecting bias in meta-analysis. Int J Evid Based Healthc. 2018;16(4):195–203. doi: 10.1097/XEB.0000000000000141

27. Duval S, Tweedie R. Trim and Fill: A Simple Funnel-Plot-Based Method of Testing and Adjusting for Publication Bias in Meta-Analysi. Biometrics. 2000;56(2):455–63. doi: 10.1111/j.0006-341x.2000.00455.x

28. Lakkireddy M, Goud Gadiga S, Malathi RD, Latha Karra M, Murthy Raju IP, Ragini, et al. Effect of Short Term High Dose Oral Vitamin D Therapy on the Inflammatory Markers in Patients with COVID 19 Disease. Archives of Clinical and Biomedical Research. 2022;06(04). doi: 10.26502/acbr.50170273

29. Soliman AR, Abdelaziz TS, Fathy A. Impact of Vitamin D Therapy on the Progress COVID-19: Six Weeks Follow-Up Study of Vitamin D Deficient Elderly Diabetes Patients. Proceedings of Singapore Healthcare. 2021;31. doi: 10.1177/20101058211041405

30. Sanchez-Zuno GA, Gonzalez-Estevez G, Matuz-Flores MG, Macedo-Ojeda G, Hernandez-Bello J, Mora-Mora JC, et al. Vitamin D Levels in COVID-19 Outpatients from Western Mexico: Clinical Correlation and Effect of Its Supplementation. J Clin Med. 2021;10(11). doi: 10.3390/jcm10112378

31. Maghbooli Z, Sahraian MA, Jamalimoghadamsiahkali S, Asadi A, Zarei A, Zendehdel A, et al. Treatment With 25-Hydroxyvitamin D(3) (Calcifediol) Is Associated With a Reduction in the Blood Neutrophil-to-Lymphocyte Ratio Marker of Disease Severity in Hospitalized Patients With COVID-19: A Pilot Multicenter, Randomized, Placebo-Controlled, Double-Blinded Clinical Trial. Endocr Pract. 2021;27(12):1242–51. doi: 10.1016/j.eprac.2021.09.016

32. Murai IH, Fernandes AL, Antonangelo L, Gualano B, Pereira RMR. Effect of a Single High-Dose Vitamin D3 on the Length of Hospital Stay of Severely 25-Hydroxyvitamin D-Deficient Patients with COVID-19. Clinics (Sao Paulo). 2021;76:e3549. doi: 10.6061/clinics/2021/e3549

33. De Niet S, Trémège M, Coffiner M, Rousseau AF, Calmes D, Frix AN, et al. Positive Effects of Vitamin D Supplementation in Patients Hospitalized for COVID-19: A Randomized, Double-Blind, Placebo-Controlled Trial. Nutrients. 2022;14(15). doi: 10.3390/nu14153048

34. Elamir YM, Amir H, Lim S, Rana YP, Lopez CG, Feliciano NV, et al. A randomized pilot study using calcitriol in hospitalized COVID-19 patients. Bone. 2022;154:116175. doi: 10.1016/j.bone.2021.116175

35. Karonova TL, Golovatyuk KA, Kudryavtsev IV, Chernikova AT, Mikhaylova AA, Aquino AD, et al. Effect of Cholecalciferol Supplementation on the Clinical Features and Inflammatory Markers in Hospitalized COVID-19 Patients: A Randomized, Open-Label, Single-Center Study. Nutrients. 2022;14(13). doi: 10.3390/nu14132602

36. Mariani J, Antonietti L, Tajer C, Ferder L, Inserra F, Sanchez Cunto M, et al. High-dose vitamin D versus placebo to prevent complications in COVID-19 patients: Multicentre randomized controlled clinical trial. PLoS One. 2022;17(5):e0267918. doi: 10.1371/journal.pone.0267918

37. Entrenas Castillo M, Entrenas Costa LM, Vaquero Barrios JM, Alcalá Díaz JF, López Miranda J, Bouillon R, et al. Effect of calcifediol treatment and best available therapy versus best available therapy on intensive care unit admission and mortality among patients hospitalized for COVID-19: A pilot randomized clinical study. J Steroid Biochem Mol Biol. 2020;203:105751. doi: 10.1016/j.jsbmb.2020.105751

38. Annweiler C, Beaudenon M, Gautier J, Gonsard J, Boucher S, Chapelet G, et al. High-dose versus standard-dose vitamin D supplementation in older adults with COVID-19 (COVIT-TRIAL): A multicenter, open-label, randomized controlled superiority trial. PLoS Med. 2022;19(5):e1003999. doi: 10.1371/journal.pmed.1003999

39. Sabico S, Enani MA, Sheshah E, Aljohani NJ, Aldisi DA, Alotaibi NH, et al. Effects of a 2-Week 5000 IU versus 1000 IU Vitamin D3 Supplementation on Recovery of Symptoms in Patients with Mild to Moderate Covid-19: A Randomized Clinical Trial. Nutrients. 2021;13(7). doi: 10.3390/nu13072170

40. Cervero M, López-Wolf D, Casado G, Novella-Mena M, Ryan-Murua P, Taboada-Martínez ML, et al. Beneficial Effect of Short-Term Supplementation of High Dose of Vitamin D(3) in Hospitalized Patients With COVID-19: A Multicenter, Single-Blinded, Prospective Randomized Pilot Clinical Trial. Front Pharmacol. 2022;13:863587. doi: 10.3389/fphar.2022.863587

41. Smaha J, Kužma M, Jackuliak P, Nachtmann S, Max F, Tibenská E, et al. Serum 25-hydroxyvitamin D Concentration Significantly Decreases in Patients with COVID-19 Pneumonia during the First 48 Hours after Hospital Admission. Nutrients. 2022;14(12). doi: 10.3390/nu14122362

42. Villasis-Keever MA, López-Alarcón MG, Miranda-Novales G, Zurita-Cruz JN, Barrada-Vázquez AS, González-Ibarra J, et al. Efficacy and Safety of Vitamin D Supplementation to Prevent COVID-19 in Frontline Healthcare Workers. A Randomized Clinical Trial. Arch Med Res. 2022;53(4):423–30. doi: 10.1016/j.arcmed.2022.04.003

43. Abroug H, Maatouk A, Bennasrallah C, Dhouib W, Ben Fredj M, Zemni I, et al. Effect of vitamin D supplementation versus placebo on recovery delay among COVID-19 Tunisian patients: a randomized-controlled clinical trial. Trials. 2023;24(1):123. doi: 10.1186/s13063-023-07114-5

44. Bishop CW, Ashfaq A, Melnick JZ, Vazquez-Escarpanter E, Fialkow JA, Strugnell SA, et al. REsCue trial: Randomized controlled clinical trial with extended-release calcifediol in symptomatic COVID-19 outpatients. Nutrition. 2023;107:111899. doi: 10.1016/j.nut.2022.111899

45. Rastogi A, Bhansali A, Khare N, Suri V, Yaddanapudi N, Sachdeva N, et al. Short term, high-dose vitamin D supplementation for COVID-19 disease: a randomised, placebo-controlled, study (SHADE study). Postgrad Med J. 2022;98(1156):87-90. doi: 10.1136/postgradmedj-2020-139065

46. Jevalikar G, Mithal A, Singh A, Sharma R, Farooqui KJ, Mahendru S, et al. Lack of association of baseline 25-hydroxyvitamin D levels with disease severity and mortality in Indian patients hospitalized for COVID-19. Sci Rep. 2021;11(1):6258. doi: 10.1038/s41598-021-85809-y

47. Nogues X, Ovejero D, Pineda-Moncusí M, Bouillon R, Arenas D, Pascual J, et al. Calcifediol Treatment and COVID-19-Related Outcomes. J Clin Endocrinol Metab. 2021;106(10):e4017–e27. doi: 10.1210/clinem/dgab405

48. Amrein K, Schnedl C, Holl A, Riedl R, Christopher KB, Pachler C, et al. Effect of high-dose vitamin D3on hospital length of stay in critically ill patients with vitamin D deficiency: The VITdAL-ICU randomized clinical trial. JAMA - Journal of the American Medical Association. 2014;312(15):1520–30. doi: 10.1001/jama.2014.13204

49. Ginde AA, Brower RG, Caterino JM, Finck L, Banner-Goodspeed VM, Grissom CK, et al. Early High-Dose Vitamin D(3) for Critically Ill, Vitamin D-Deficient Patients. N Engl J Med. 2019;381(26):2529–40. doi: 10.1056/NEJMoa1911124

50. Else H. How a torrent of COVID science changed research publishing - in seven charts. Nature. 2020;588(7839):553. doi: 10.1038/d41586-020-03564-y

51. Sterne JA, Sutton AJ, Ioannidis JP, Terrin N, Jones DR, Lau J, et al. Recommendations for examining and interpreting funnel plot asymmetry in meta-analyses of randomised controlled trials. BMJ. 2011;343:d4002. doi: 10.1136/bmj.d4002

52. Borenstein M. Common mistakes in meta-analysis. New Jersey: Biostat, Inc.; 2019.

53. Lau J, Ioannidis JP, Terrin N, Schmid CH, Olkin I. The case of the misleading funnel plot. Bmj. 2006;333(7568):597-600. doi: 10.1136/bmj.333.7568.597

54. Schwab S, Kreiliger G, Held L. Assessing treatment effects and publication bias across different specialties in medicine: a meta-epidemiological study. BMJ Open. 2021;11(9):e045942. doi: 10.1136/bmjopen-2020-045942

55. Ioannidis JP, Greenland S, Hlatky MA, Khoury MJ, Macleod MR, Moher D, et al. Increasing value and reducing waste in research design, conduct, and analysis. Lancet. 2014;383(9912):166-75. doi: 10.1016/s0140-6736(13)62227-8

56. Kjaergard LL, Villumsen J, Gluud C. Reported methodologic quality and discrepancies between large and small randomized trials in meta-analyses. Ann Intern Med. 2001;135(11):982–9. doi: 10.7326/0003-4819-135-11-200112040-00010

57. Zhang Z, Xu X, Ni H. Small studies may overestimate the effect sizes in critical care meta-analyses: a meta-epidemiological study. Crit Care. 2013;17(1):R2. doi: 10.1186/cc11919

58. Hopefl R, Ben-Eltriki M, Deb S. Association Between Vitamin D Levels and Inflammatory Markers in COVID-19 Patients: A Meta-Analysis of Observational Studies. J Pharm Pharm Sci. 2022;25:124–36. doi: 10.18433/jpps32518

59. Antonelli M, Kushner I. Low Serum Levels of 25-Hydroxyvitamin D Accompany Severe COVID-19 Because it is a Negative Acute Phase Reactant. Am J Med Sci. 2021;362(3):333–5. doi: 10.1016/j.amjms.2021.06.005

60. Binkley N, Coursin D, Krueger D, Iglar P, Heiner J, Illgen R, et al. Surgery alters parameters of vitamin D status and other laboratory results. Osteoporos Int. 2017;28(3):1013–20. doi: 10.1007/s00198-016-3819-9

61. Silva MC, Furlanetto TW. Does serum 25-hydroxyvitamin D decrease during acute-phase response? A systematic review. Nutr Res. 2015;35(2):91–6. doi: 10.1016/j.nutres.2014.12.008

62. Autier P, Boniol M, Pizot C, Mullie P. Vitamin D status and ill health: a systematic review. The Lancet Diabetes & Endocrinology. 2014;2(1):76–89. doi: 10.1016/s2213-8587(13)70165-7

63. Looker AC. Serum 25-hydroxyvitamin D and risk of major osteoporotic fractures in older U.S. adults. J Bone Miner Res. 2013;28(5):997–1006. doi: 10.1002/jbmr.1828

64. Scragg R, Stewart AW, Waayer D, Lawes CM, Toop L, Sluyter J, et al. Effect of Monthly High-Dose Vitamin D Supplementation on Cardiovascular Disease in the Vitamin D Assessment Study : A Randomized Clinical Trial. JAMA Cardiol. 2017. doi: 10.1001/jamacardio.2017.0175

65. Pittas AG, Dawson-Hughes B, Sheehan P, Ware JH, Knowler WC, Aroda VR, et al. Vitamin D Supplementation and Prevention of Type 2 Diabetes. N Engl J Med. 2019;381(6):520–30. doi: 10.1056/NEJMoa1900906

66. Yao P, Bennett D, Mafham M, Lin X, Chen Z, Armitage J, et al. Vitamin D and Calcium for the Prevention of Fracture: A Systematic Review and Meta-analysis. JAMA Netw Open. 2019;2(12):e1917789. doi: 10.1001/jamanetworkopen.2019.17789

67. Autier P, Mullie P, Macacu A, Dragomir M, Boniol M, Coppens K, et al. Effect of vitamin D supplementation on non-skeletal disorders: a systematic review of meta-analyses and randomised trials. The Lancet Diabetes & Endocrinology. 2017;5(12):986–1004. doi: 10.1016/s2213-8587(17)30357-1

68. Subramanian S, Griffin G, Hewison M, Hopkin J, Kenny RA, Laird E, et al. Vitamin D and COVID-19-Revisited. J Intern Med. 2022;292(4):604–26. doi: 10.1111/joim.13536

69. Martineau AR. Vitamin D in the prevention or treatment of COVID-19. Proc Nutr Soc. 2022:1–8. doi: 10.1017/s0029665122002798

